# Nasal colonizing vancomycin-resistant and intermediate *Staphylococcus aureus* among admitted patients

**DOI:** 10.1101/2024.08.12.24311719

**Authors:** Biniyam Kijineh Mengistu, Tsegaye Alemayehu, Techilo Habtemariam Mengesha, Musa Mohammed Ali

## Abstract

**Background:** *Staphylococcus aureus* colonizing the nasal cavity is a potential source of infections. Vancomycin is a mainstay for treating invasive infections caused by penicillin and methicillin-resistant *S. aureus* (MRSA). Some reports indicate the emergence of vancomycin-resistant *S. aureus* (VRSA) making it a high-priority pathogen that needs attention. There is a limited report on the epidemiology of VRSA and vancomycin-intermediate *S. aureus* (VISA) from the Sidama regional state.

**Objective:** The objective of this study was to determine VRSA and VISA among *S. aureus* colonizing patients admitted at Hawassa University Comprehensive Specialized Hospital (HUCSH), associated factors, and antimicrobial susceptibility profile.

**Methods:** A hospital-based prospective cross-sectional study was conducted from April to June 2023. Socio-demographic and clinical data were collected using an interviewer-administered questionnaire. Nasal swabs were collected from 378 admitted patients. Identification of *S. aureus* was made using standard bacteriological methods. VRSA was determined by the Epsilometer test (E-test). The antimicrobial susceptibility profile was determined according to the Kirby-Bauer disk diffusion method. Data was analyzed using SPSS version 22. A *p*<0.05 was taken as a cut point to determine a statistically significant association.

**Results:** Out of the total 92 *S. aureus* isolated 12 (13.04%), 27(29.3%), 15(16.3%) were VRSA, VISA, and MRSA respectively. The carriage rate of VRSA and VISA among admitted patients were 12(3.2%) with 95% CI: 1.7%−5.5% and 27(7.14%) with 95% CI: 4.8%−10.2% respectively. The overall nasal carriage rate of *S. aureus* and MRSA was 92(24.3%) with 95% CI: 20.1%−29% and 15(3.97%) with 95% CI: 2.2%−6.5% respectively. Of the VRSA isolates, 11(91.7%) were susceptible to tigecycline. Forty (43.5%) of *S. aureus* were positive for inducible clindamycin resistance. Participants with a history of hospitalization at the intensive care unit were 37 times more likely to be colonized with VRSA (*p*=0.001). Participants who have domestic animals were 22 times more likely to be colonized with VRSA (*p*=0.021).

**Conclusions:** This study indicated a high proportion of VRSA and VISA among *S. aureus* isolated from hospitalized patients in the study area. More than 80% of VRSA were susceptible to tigecycline. History of hospitalization at the intensive care unit and having domestic animals at home could increase the odds of VRSA colonization.

## Background

Antimicrobial resistance (AMR) is one of the most important public health problems in the world disproportionately affecting low-income countries (1). Worldwide dissemination of drug-resistant bacteria with few developments of antimicrobial agents is a formidable challenge that the physicians of today are facing (1).

*Staphylococcus aureus* is among potential pathogenic bacteria with diverse virulence factors and the potential to develop resistance to commonly used antimicrobial agents. *S. aureus* are colonizer of the skin and upper respiratory tract without causing diseases. If there are underlying conditions it can cause community and hospital-acquired infections such as soft tissue infections, bacteremia, endocarditis, pneumonia, meningitis, and osteomyelitis (2).

*S. aureus* is a frequent inhabitant of the human upper respiratory tract and the skin (3); however, the carriage rate varies based on geographic location, age, gender, and ethnicity (4). The nasal carriage of *S. aureus* has been identified as a risk factor for community-acquired and nosocomial infections (5).

*S. aureus* has developed resistance to penicillin and methicillin over time, which makes infection caused by *S. aureus* difficult to treat (6). Vancomycin, a glycopeptide antibiotic, has been used as a last-line antibiotic for treating life-threatening infections caused by Methicillin-resistant *S. aureus* (MRSA) (7). This option is being challenged by the emergence and spread of *S. aureus* which are completely resistant to vancomycin or reduced susceptibility to vancomycin (7, 8).

*S. aureus* strains exhibiting reduced susceptibility to vancomycin, vancomycin intermediate-resistant *S. aureus* (VISA), were discovered in the 1990s. *S. aureus* isolates with complete resistance to vancomycin, vancomycin-resistant *S. aureus* (VRSA), was first reported in the United States in 2002. The burden of VISA is higher than VRSA; it is associated with persistent infections, vancomycin treatment failure, and poor clinical outcomes (7, 8). VRSA was among the list of antibiotic-resistant microorganisms identified by the World Health Organization (WHO) in 2017 for which new antibiotics should be developed (9). Furthermore, VRSA remained in the 2019 WHO updated list as a high-priority pathogen (10).

Infections caused by VRSA have been associated with high morbidity and mortality rates due to the limited availability of effective drugs, particularly in low-income countries. In Ethiopia, the prevalence of drug-resistant bacteria has been increasing from time to time (11). The prevalence of VRSA reported from other parts of Ethiopia (12, 14) is mostly based on the disc diffusion method. Some studies from Ethiopia used E-test and Micro-dilution methods for the determination of VRSA (15, 16). In addition to searching for alternative treatments for infection caused by VRSA, it is important to monitor the epidemiology of VRSA. Considering the scarcity of data in the southern parts of Ethiopia and the disparity in methods used for the detection of VRSA, we primarily envisaged determining the nasal carriage rate of VRSA among admitted patients, its predictors, and antimicrobial susceptibility profile. We also determined the proportion of VISA, MRSA, and ICR *S. aureus*.

## Methods

### Study design and study population

A hospital-based prospective cross-sectional study was conducted among patients admitted to Hawassa University Comprehensive Specialized Hospital (HUCSH) from April to June 2023. The hospital is located in Hawassa city, which is the capital city of Sidama regional state, Ethiopia. HUCSH is a referral hospital that provides health-related services for people coming from the Sidama regional state and other neighboring regional states. It has 284 functional beds and 12 wards such as surgical ward, medical ward, ophthalmology ward, neurology ward, gynecology ward, orthopedics ward, pediatrics ward, pedi-surgery ward, oncology ward, adult and neonatal intensive care units, and trauma ward. We have included six wards for the present study based on the patient flow.

In this study, we included patients who were admitted to the HUCSH during the study period for treatment. Patients who were critically sick and had nasal bleeding were excluded from the study. The sample size was determined based on a single sample size estimation formula. We have considered the prevalence of VRSA (4.1%) which was reported from Debre Markos, Ethiopia (17). By assuming a 5% for non-response rate, the sample size was 378. A total of 2,222 patients were admitted from September to December. The sample size was proportionally allocated to seven wards by multiplying, 378/2222 (0.17), with a number of patients admitted at each ward between September to December 2022 (Figure 1). A systematic random sampling technique was used to select study participants. The k-value was determined based on the total number of patients allocated to each ward. The first participant was selected using the lottery method.

**Figure 1.**
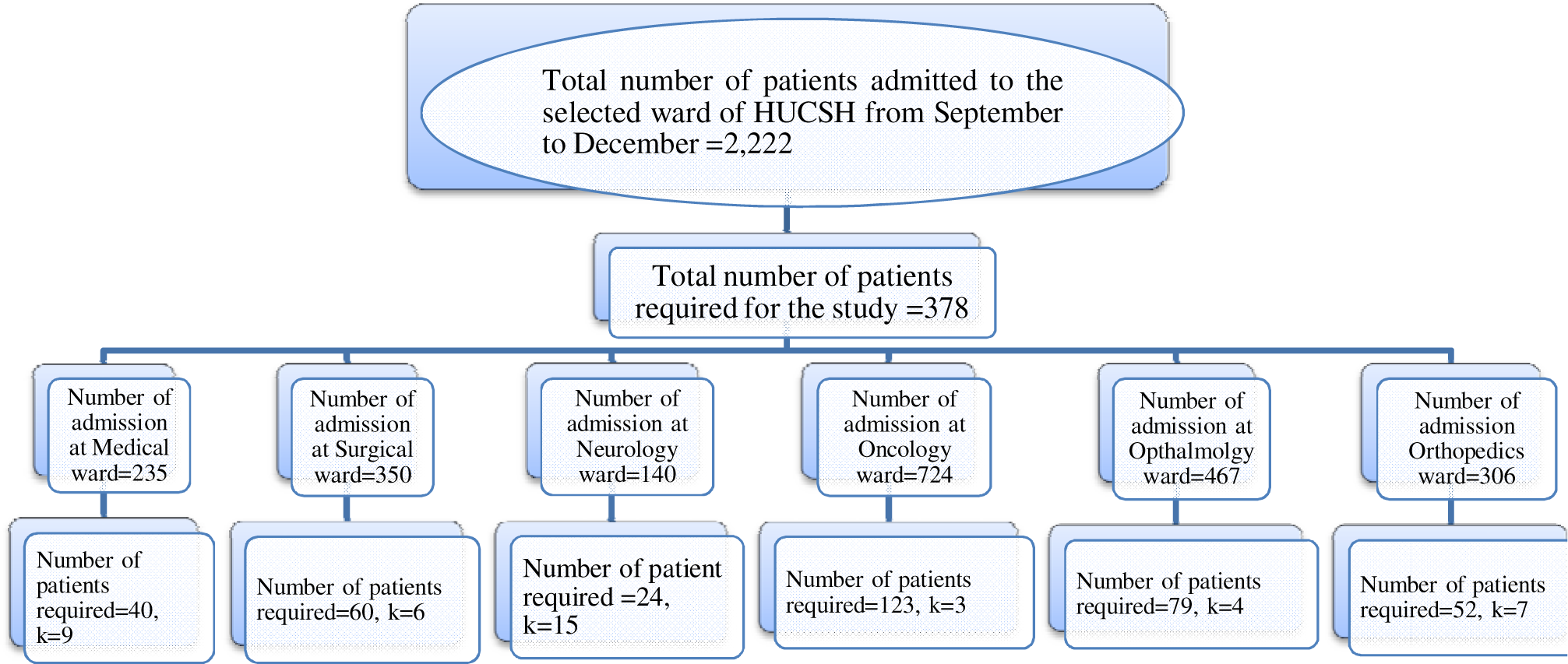
Total number of patients admitted to selected wards of HUCSH from September to December 2022 and proportional allocation of participant at each ward.

### Data Collection

A structured questionnaire prepared on kobo-collect version 2023.1.2 (United States) was used to collect socio-demographic and clinical data. The questionnaire was prepared after reviewing similar published articles (5, 12, 13, 19). To ensure consistency, the questionnaire was first prepared in English, translated into Amharic, and then translated back into English. The data was collected using the Amharic version of the questionnaire. The questionnaire was pretested in a population representing 5% (n=19) of the sample size at Adare General Hospital, Hawassa, Ethiopia.

The anterior nostrils were swabbed aseptically using sterile cotton soaked in saline and placed in a tube containing normal saline. The swabs were inoculated onto mannitol salt agar (MSA) (Oxoid, Hampshire UK) and incubated at 37°C for 24 hours in an aerobic environment. After 24 hours of incubation, MSA plates were inspected for visible growth of bacteria. In cases where there was no visible growth of bacteria, plates were re-incubated for an additional 24 hours. For bacterial growth with yellow colonies on MSA, Gram staining was performed. Gram-positive cocci bacteria in a cluster on microscopic examination were tested using catalase, coagulase, and modified oxidase test. Bacteria were considered as *S. aureus* if it appeared yellow on MSA, catalase positive, coagulase-positive and modified oxidase test negative. We used a modified oxidase test to differentiate *S. aureus* from *Micrococcus* which may appear yellow on MSA and can wrongly be considered as *S. aureus.* Confirmed *S. aureus* isolates were tested for VRSA, MRSA, and antimicrobial susceptibility testing. Before use, the sterility of all culture media used in this study was checked by incubating 5% of each batch of the culture media without inoculation. For performance testing of Gram staining, culture media, biochemical tests, and antibiotics reference strains were used.

### Detection of vancomycin resistant *S. aureus*

The pure isolate of *S. aureus* was suspended in a phosphate buffer solution until it matched the McFarland 0.5 standard. The suspension was inoculated over the surface of Muller Hinton Agar (MHA) uniformly using a sterile cotton swab dipped into a suspension. Vancomycin E-test (bioMérieux SA, Marcy-1’Étoile, France) was applied to MHA in which *S. aureus* was inoculated and incubated for 24 hours aerobically at 37°C. The reading was interpreted according to Clinical Laboratory Standard Institute (CLSI) guidelines as susceptible, intermediate, and resistant (18). Minimum inhibitory concentrations of ≤2 μg/ml, 4–8μg/ml, and ≥16μg/ml were interpreted as susceptible (S), intermediate (I), and resistant (R) respectively.

To detect MRSA, *S. aureus* suspension was prepared and inoculated onto MHA as mentioned above; a cefoxitin (30μg) antibiotic was applied and incubated for 18 hours in ambient air at 35°C. Zone of inhibition <21 mm was interpreted as MRSA. Inducible clindamycin resistance (ICR) was identified using a D-test which involves the application of erythromycin (E-15 μg) and clindamycin (CD-2 μg) discs spaced 15−26 mm on MHA as per CLSI guidelines (18). Flattening of the zone of inhibition adjacent to the erythromycin disc and cloudy growth within the zone of inhibition around clindamycin indicates ICR.

### Antibiotic susceptibility testing

The disc diffusion method was used to determine the antibiotic susceptibility profile of S*. aureus* as described in CLSI guidelines (18). The suspension of *S. aureus* was prepared and inoculated onto MHA as described above. The antibiotics used for disc diffusion assays include cefoxitin (30µg), erythromycin (15µg), trimethoprim-sulfamethoxazole (1.25/23.7, 5µg), ciprofloxacin (5µg), chloramphenicol (30µg), gentamicin (10µg), clindamycin (2µg), azithromycin (15µg), tigecycline (15µg), aztreonam (30µg), doxycycline (30µg), penicillin G (10 unit). MHA was incubated at 35−37°C for 24 hours. The zones of inhibition were measured using a caliper, and interpreted as susceptible (S), intermediate (I), or resistant (R).

### Data Analysis and Interpretation

Data was cleaned, transformed, coded, and analyzed using SPSS version 22. Data was presented in frequency and percent. The association between independent and dependent variables was determined using the binary logistic regression model; variables with *p*≤ 0.25 in bivariate analysis were included in the multivariable logistic regression model to identify variables that predict VRSA colonization. All independent variables with a *p*≤ 0.05 were considered statistically significant.

## Results

### Socio demographic and clinical characteristics

A total of 378 admitted patients at HUCSH were included in the study with a 100% response rate. The majority (n=215; 56.9%) of the study participants were males. The mean age of study participants was 44.5 with a standard deviation (SD) of 19.2 years. Most of the study participants (n=160; 42.3%) were from Sidama Regional State. More than half of the participants were on IV or catheter while 126 (33.3%) reported repeated use of antibiotics (Table 1).

**Table 1.**
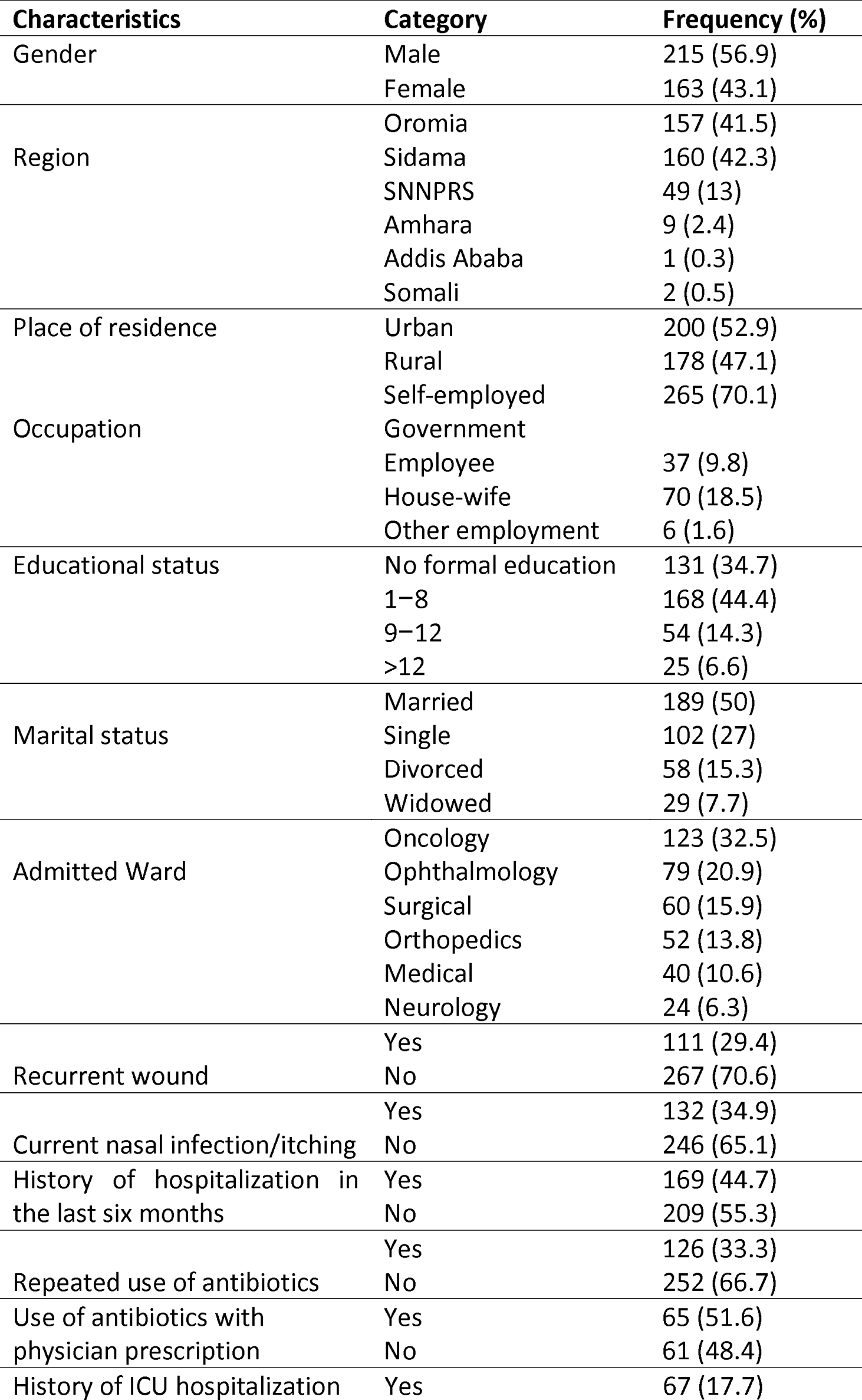
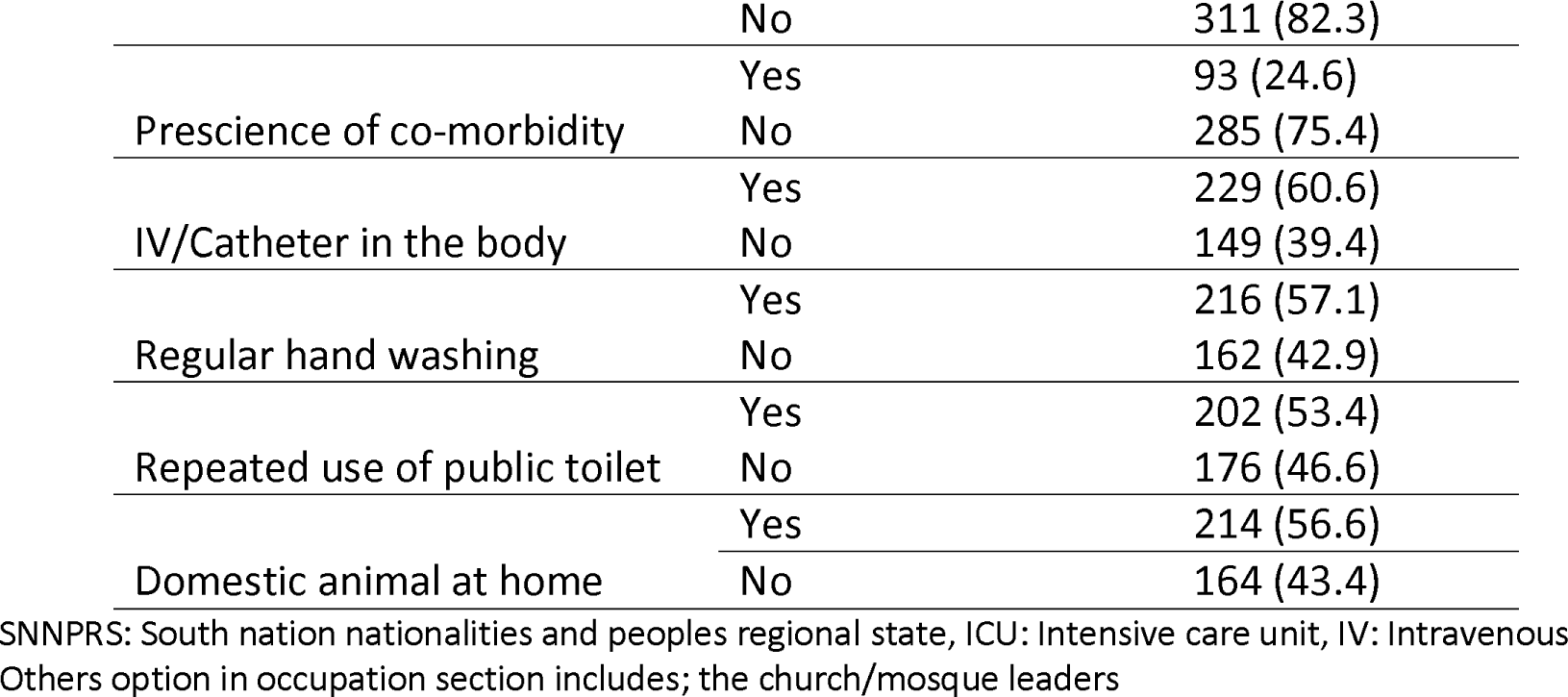
Socio-demographic and clinical characteristics of admitted patients at Hawassa HUCSH, Hawassa, Ethiopia, April to June 2023 (N=378).

### Prevalence of *S. aureus*, VRSA, and MRSA

Out of the total *S. aureus*isolated, 12(13.04%) 95% CI: 6.9%−21.7% and 27(29.3%) 95% CI; 20.3%−39.8% were VRSA and VISA. 15(16.3%) 95% CI: 9.4%-25.5% of *S. aureus* were MRSA. Out of 15 MRSA, 3 were vancomycin resistant. Out of the total *S. aureus* isolated, 21 (22.83%) were susceptible to both erythromycin and clindamycin whereas 31(33.7%) were resistant to both erythromycin and clindamycin indicating constitutive resistance (cMLSB). Forty (43.5%) of *S. aureus* were resistant to erythromycin and susceptible to clindamycin indicating ICR (iMLSB). The prevalence of VRSA, VISA, MRSA, and overall *S. aureus* colonization rate among admitted patients were 12(3.2%) 95% CI: 1.7%−5.5%, 27(7.14%) 95%CI: 4.8%−10.2%, 15(3.97%) 95%CI: 2.2%−6.5%, and 92(24.3%) 95% CI: 20.1%−29% respectively.

### Factors associated with VRSA

In bivariate logistic regression analysis, recurrent wound (*p*=0.04, COR=3.69(95% CI:1.06−12.9)), nasal infection (*p*=0.024, COR=4.4(95%CI:1.21−15.9)), history of hospitalization (*p*=0.033, COR=4.5(95% CI: 1.13−17.9)), repeated use of antibiotics (*p*=0.04, COR=3.93(95%CI:1.08−14.2)), history of hospitalization at ICU (*p*<0.0001,COR= 31.4(95% CI:6.05−162.7)) and having domestic animal at home (*p*=0.019,COR=12.16(95%CI: 1.49−98.6)) were significantly associated with VRSA colonization. Variables with *p*< 0.25 in the bivariate logistic regression model were included in multivariable logistic analysis. History of hospitalization at ICU (*p*=0.001, AOR=37.2(95% CI: 4.32−320.8)) and having a domestic animal at home (*p*=0.021, AOR=22.37(95% CI: 1.59−313.1)) were significantly associated with VRSA colonization in multivariable logistic regression analysis (Table 2).

**Table 2.**
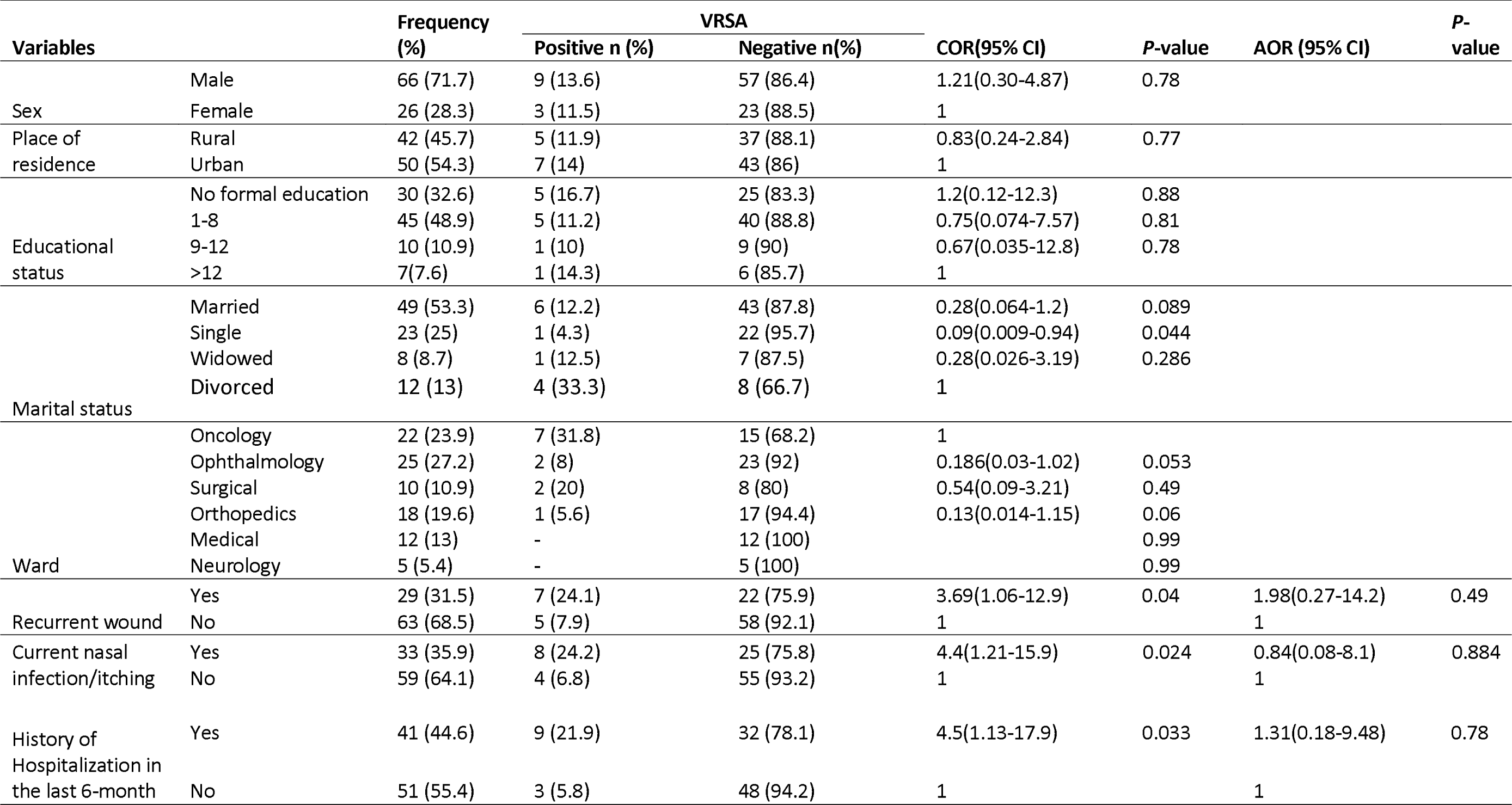
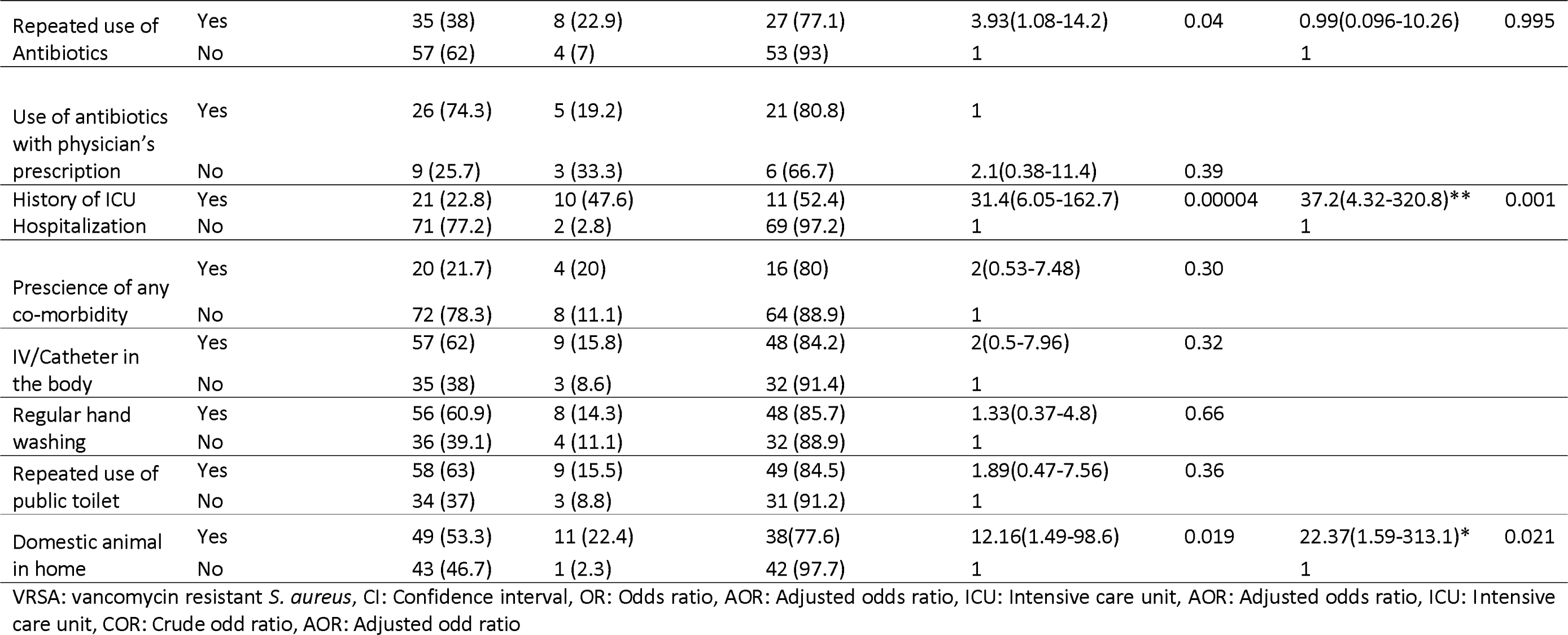
Factors associated with VRSA carriage among patients admitted at HUCSH, Hawassa, Ethiopia, April to June 2023 (N=92).

| Variables |  | Frequency (%) | VRSA |  | COR(95% CI) | P-value | AOR (95% CI) | P-value |
| --- | --- | --- | --- | --- | --- | --- | --- | --- |
|  |  |  | Positive n (%) | Negative n(%) |  |  |  |  |
| Sex | Male | 66 (71.7) | 9 (13.6) | 57 (86.4) | 1.21(0.30-4.87) | 0.78 |  |  |
|  | Female | 26 (28.3) | 3 (11.5) | 23 (88.5) | 1 |  |  |  |
| Place of residence | Rural | 42 (45.7) | 5 (11.9) | 37 (88.1) | 0.83(0.24-2.84) | 0.77 |  |  |
|  | Urban | 50 (54.3) | 7 (14) | 43 (86) | 1 |  |  |  |
| Educational status | No formal education | 30 (32.6) | 5 (16.7) | 25 (83.3) | 1.2(0.12-12.3) | 0.88 |  |  |
|  | 1-8 | 45 (48.9) | 5 (11.2) | 40 (88.8) | 0.75(0.074-7.57) | 0.81 |  |  |
|  | 9-12 | 10 (10.9) | 1 (10) | 9 (90) | 0.67(0.035-12.8) | 0.78 |  |  |
|  | >12 | 7(7.6) | 1 (14.3) | 6 (85.7) | 1 |  |  |  |
| Marital status | Married | 49 (53.3) | 6 (12.2) | 43 (87.8) | 0.28(0.064-1.2) | 0.089 |  |  |
|  | Single | 23 (25) | 1 (4.3) | 22 (95.7) | 0.09(0.009-0.94) | 0.044 |  |  |
|  | Widowed | 8 (8.7) | 1 (12.5) | 7 (87.5) | 0.28(0.026-3.19) | 0.286 |  |  |
|  | Divorced | 12 (13) | 4 (33.3) | 8 (66.7) | 1 |  |  |  |
| Ward | Oncology | 22 (23.9) | 7 (31.8) | 15 (68.2) | 1 |  |  |  |
|  | Ophthalmology | 25 (27.2) | 2 (8) | 23 (92) | 0.186(0.03-1.02) | 0.053 |  |  |
|  | Surgical | 10 (10.9) | 2 (20) | 8 (80) | 0.54(0.09-3.21) | 0.49 |  |  |
|  | Orthopedics | 18 (19.6) | 1 (5.6) | 17 (94.4) | 0.13(0.014-1.15) | 0.06 |  |  |
|  | Medical | 12 (13) | - | 12 (100) |  | 0.99 |  |  |
|  | Neurology | 5 (5.4) | - | 5 (100) |  | 0.99 |  |  |
| Recurrent wound | Yes | 29 (31.5) | 7 (24.1) | 22 (75.9) | 3.69(1.06-12.9) | 0.04 | 1.98(0.27-14.2) | 0.49 |
|  | No | 63 (68.5) | 5 (7.9) | 58 (92.1) | 1 |  | 1 |  |
| Current nasal infection/itching | Yes | 33 (35.9) | 8 (24.2) | 25 (75.8) | 4.4(1.21-15.9) | 0.024 | 0.84(0.08-8.1) | 0.884 |
|  | No | 59 (64.1) | 4 (6.8) | 55 (93.2) | 1 |  | 1 |  |
| History of Hospitalization in the last 6-month | Yes | 41 (44.6) | 9 (21.9) | 32 (78.1) | 4.5(1.13-17.9) | 0.033 | 1.31(0.18-9.48) | 0.78 |
|  | No | 51 (55.4) | 3 (5.8) | 48 (94.2) | 1 |  | 1 |  |
| Repeated use of Antibiotics | Yes | 35 (38) | 8 (22.9) | 27 (77.1) | 3.93(1.08-14.2) | 0.04 | 0.99(0.096-10.26) | 0.995 |
|  | No | 57 (62) | 4 (7) | 53 (93) | 1 |  | 1 |  |
| Use of antibiotics with physician's prescription | Yes | 26 (74.3) | 5 (19.2) | 21 (80.8) | 1 |  |  |  |
|  | No | 9 (25.7) | 3 (33.3) | 6 (66.7) | 2.1(0.38-11.4) | 0.39 |  |  |
| History of ICU Hospitalization | Yes | 21 (22.8) | 10 (47.6) | 11 (52.4) | 31.4(6.05-162.7) | 0.00004 | 37.2(4.32-320.8)** | 0.001 |
|  | No | 71 (77.2) | 2 (2.8) | 69 (97.2) | 1 |  | 1 |  |
| Prescience of any co-morbidity | Yes | 20 (21.7) | 4 (20) | 16 (80) | 2(0.53-7.48) | 0.30 |  |  |
|  | No | 72 (78.3) | 8 (11.1) | 64 (88.9) | 1 |  |  |  |
| IV/Catheter in the body | Yes | 57 (62) | 9 (15.8) | 48 (84.2) | 2(0.5-7.96) | 0.32 |  |  |
|  | No | 35 (38) | 3 (8.6) | 32 (91.4) | 1 |  |  |  |
| Regular hand washing | Yes | 56 (60.9) | 8 (14.3) | 48 (85.7) | 1.33(0.37-4.8) | 0.66 |  |  |
|  | No | 36 (39.1) | 4 (11.1) | 32 (88.9) | 1 |  |  |  |
| Repeated use of public toilet | Yes | 58 (63) | 9 (15.5) | 49 (84.5) | 1.89(0.47-7.56) | 0.36 |  |  |
|  | No | 34 (37) | 3 (8.8) | 31 (91.2) | 1 |  |  |  |
| Domestic animal in home | Yes | 49 (53.3) | 11 (22.4) | 38(77.6) | 12.16(1.49-98.6) | 0.019 | 22.37(1.59-313.1)* | 0.021 |
|  | No | 43 (46.7) | 1 (2.3) | 42 (97.7) | 1 |  | 1 |  |

VRSA: vancomycin resistant *S. aureus*, CI: Confidence interval, OR: Odds ratio, AOR: Adjusted odds ratio, ICU: Intensive care unit, AOR: Adjusted odds ratio, ICU: Intensive care unit, COR: Crude odd ratio, AOR: Adjusted odd ratio

In bivariate analysis gender (*p*=0.001,COR=2.3 (95%CI:1.38−3.8)), repeated use of antibiotics with physician’s prescription (*p*=0.002, COR=0.26(95%CI:0.11−0.62)), and repeated use of public toilet (*p*=0.035, COR=1.68(95% CI:1.04−2.73)) were significantly associated with overall *S. aureus* colonization rate while in multivariable analysis gender (*p*=0.008, AOR=3.67(95%CI:1.4−9.6)), repeated use of antibiotics with physician’s prescription (*p*= 0.009, AOR=0.28(95% CI: 0.11−0.73)) and history of hospitalization at ICU (*p*=0.023, AOR=3.34(95% CI: 1.2−9.46)) were significantly associated with overall *S. aureus* colonization rate (Supplement table 1).

In bivariate logistic analysis, history of ICU hospitalization (*p =* 0.004, COR = 5.63(95% CI: 1.73−18.2)) showed significant association while in multivariate analysis, gender (*p =* 0.018, AOR = 0.16(95% CI: 0.035−0.73)) and history of ICU hospitalization (*p =* 0.008, AOR = 8.06(95% CI: 1.72−37.6)) were significantly associated with MRSA colonization (Supplement table 2).

### Antimicrobial susceptibility profile of VRSA

Out of the total *S. aureus* tested, 92(100%), 76(82.6%), 68(73.9%), and 66(71.7%) were resistant to aztreonam, penicillin G, sulfamethoxazole-trimethoprim (SXT), and azithromycin respectively on the other hand 88(95.7%), 82(89.1%), 77(83.7%) and 71(77.2%) of *S. aureus* were susceptible to tigecycline, doxycycline, cefoxitin, and chloramphenicol respectively (Table 3).

**Table 3.** Antimicrobial susceptibility profile of *S. aureus* and VRSA recovered from patients admitted at HUCSH, Hawassa, Ethiopia. April to June 2023.

| Antimicrobials | *S. aureus* n=92 |  |  | VRSA n=12 |  |  |
| --- | --- | --- | --- | --- | --- | --- |
|  | S, n (%) | I, n (%) | R, n (%) | S, n (%) | I, n (%) | R, n (%) |
| AZM | 23 (25) | 3 (3.3) | 66 (71.7) | 1 (8.3) | 0 | 11 (91.7) |
| CD | 68 (73.9) | 1 (1.1) | 23 (25) | 5 (41.7) | 0 | 7 (58.3) |
| TGC | 88 (95.7) | NR | 4 (4.3) | 11 (91.75) | NR | 1 (8.3) |
| AZT | 0 | 0 | 92 (100) | 0 | 0 | 12 (100) |
| CN | 56 (60.9) | 1 (1.1) | 35 (38) | 7 (58.3) | 1 (8.3) | 4 (33.3) |
| CIP | 47 (51.1) | 12 (13) | 33 (35.9) | 6 (50) | 3 (25) | 3 (25) |
| ERY | 21 (22.8) | 7 (7.6) | 64 (69.6) | 1 (8.3) | 2 (16.7) | 9 (75) |
| DOX | 82 (89.1) | 4 (4.3) | 6 (6.5) | 10 (83.3) | 1 (8.3) | 1 (8.3) |
| C | 71 (77.2) | 15 (16.3) | 6 (6.5) | 6 (50) | 3 (25) | 3 (25) |
| PEN G | 16 (17.4) | NR | 76 (82.6) | 6 (50) | NR | 6 (50) |
| SXT | 24 (26.1) | 0 | 68 (73.9) | 1 (8.3) | 0 | 11 (91.7) |
| FOX | 77 (83.7) | NR | 15 (16.3) | 9 (75) | NR | 3 (25) |

VRSA: Vancomycin resistant *S. aureus*, S: Susceptible, I: Intermediate, R: Resistant, AZM: Azithromycin, CD: Clindamycin, TGC: Tigecycline, AZT: Aztreonam, CN: Gentamicin, CIP: Ciprofloxacin, ERY: Erythromycin, DOX: Doxycycline, C: Chloramphenicol,, PEN G: Penicillin G, SXT:- Sulfamethoxazole- trimethoprim , FOX: Cefoxitin, NR: Not recommended by CLSI

Out of the total VRSA, 12(100%), 11(91.7%), 9(75%), and 7(58.3%) were resistant to aztreonam, azithromycin, clindamycin, and gentamicin respectively while 11(91.75%), 10(83.3%), 9(75%), and 7(58.3%) VRSA were susceptible to tigecycline, doxycycline, cefoxitin, and gentamicin respectively (Table 3). A high proportion of vancomycin susceptible *S. aureus* was positive for inducible clindamycin resistance while the lowest inducible clindamycin resistance was observed among VRSA (Fig 2).

**Figure 2.**
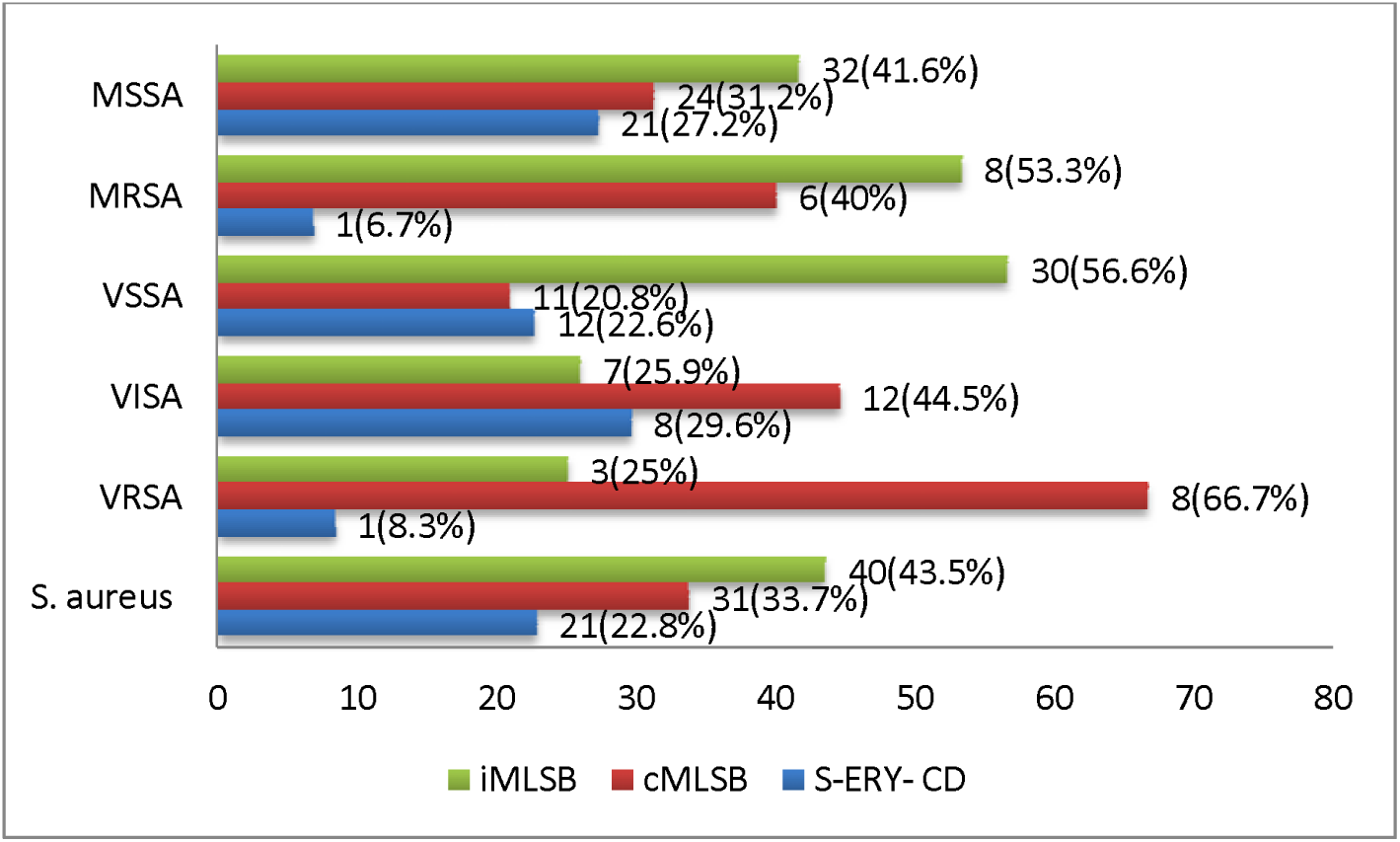
Clindamycin resistance observed across *S. aureus*, VRSA and MRSA isolated from admitted patients at HUCSH, Hawassa, Ethiopia April 05 to June 24 2023 VSSA: Vancomycin susceptible *S. aureus* VISA: Vancomycin intermediate *S. aureus*, VRSA: Vancomycin resistant *S. aureus,* S-ERY-CD: Susceptible to both Erythromycin and clindamycin, cMLSB: constitutive resistance (Resistance to both Erythromycin and Clindamycin), iMLS: inducible resistance to clindamycin, MRSA: Methicillin resistant *S. aureus,* MSSA: Methicillin susceptible *S. aureus*

## Discussion

Infections caused by VRSA are public health concerns that require attention from international and national health sectors. Treatment options for VRSA infections are limited. Assessing the epidemiology of VRSA will provide valuable insights to help combat its spread within the community. So far, a few studies in Ethiopia have identified VRSA using the minimum inhibitory concentration method.

The present study found the proportion of VRSA among *S. aureus* isolates to be 13.04%. This is consistent with studies conducted in Eritrea (15.9%) (19), Pakistan (11-15%) (20), Egypt (13.8%) (21), Ethiopia (14-15%) (6, 16, 17, 22), and Indonesia (11%) (23). However, the VRSA prevalence in this study was higher than many other studies in Addis Ababa, Ethiopia (4-7%) (14, 15, 24), the global pooled estimate (6%) (25), Yemen (5.12%) (26), and Egypt (4.5%) (27). Conversely, studies from Nigeria (44.5%) (3), Bangladesh (27.6%) (26), and Egypt (24%) (29) reported a higher VRSA prevalence than the current study. The lowest VRSA rate (4.1%) in Ethiopia was reported from Jigjiga (14).

These variations could be attributed to differences in the study population, specimen collection site, laboratory method, study period, and antibiotic usage practices. The current study did not determine the origin of VRSA in the patients, who were admitted for various medical conditions. VRSA could have been acquired in the community before admission or during hospitalization. Further investigation using molecular epidemiology is needed to elucidate the source of VRSA in these patients.

The study found that participants with a history of ICU hospitalization were more likely to be colonized with VRSA (*p*=0.001). Similar findings have been reported in previous studies by Wu et al. (25) and Shariati et al. (30). This could be attributed to the misuse of antimicrobial agents and the use of contaminated medical devices in hospital settings during patient management. Additionally, participants who had domestic animals at home were also more likely to be colonized with VRSA compared to those without domestic animals. This observation aligns with findings from other studies conducted in Ethiopia (17, 31). This may suggest that domestic animals could be a potential source of VRSA; however, the current study was not designed to definitively determine the source of VRSA infection.

The study found that all VRSA isolates were resistant to aztreonam, which is consistent with findings from a study conducted in Hyderabad (32). Additionally, over 25% of the isolates were resistant to azithromycin and sulfamethoxazole-trimethoprim, which is lower than the resistance rates reported in a study from Egypt (21). Conversely, only one isolate was resistant to tigecycline, a similar observation to a study in Pakistan (20).

The nasal carriage rate of *S. aureus* has been extensively studied, and various colonization rates have been reported across different regions and study populations (13, 33, 34, 43). The overall nasal carriage rate of *S. aureus* (24.3%) found in the present study is comparable to rates reported from Ethiopia (28.8%) (13), China (24.7%) (33), and Iran (21.3%) (43). However, it is lower than the rates observed in studies conducted in Iraq (39.4%) (34) and other parts of Ethiopia (32%-39%) (16, 35). Notably, the nasal carriage rate in this study was higher than rates reported in other regions of Ethiopia (13%-16%) (36, 37).

These variations in the nasal carriage of *S. aureus* across different regions of Ethiopia suggest the existence of other factors that may contribute to different levels of colonization rates. The differences in the isolation rates of *S. aureus* among studies could be attributed to variations in clinical characteristics, study settings, socio-demographic factors, and individual behaviors.

The study found that male participants were 3.67 times more likely to be colonized with *S. aureus*, which contradicts the findings of other studies conducted in Jimma, Ethiopia (38). This discrepancy could be attributed to behavioral differences between males and females residing in various geographic locations within Ethiopia. Interestingly, study participants who did not use antibiotics without a physician’s prescription were 72% less likely to be colonized with S*. aureus*. Additionally, participants with a history of ICU hospitalization were 3.34 times more likely to be colonized with *S. aureus*, a finding that aligns with the report by Humphreys et al. (39).

Among the 92 *S. aureus* isolates in the current study, 16.3% were resistant to cefoxitin, which is lower than the resistance rate reported in a study from Nigeria (40). However, 69.6% and 25% of the *S. aureus* isolates were resistant to erythromycin and clindamycin, respectively, which is lower than the resistance rates observed in a study conducted in Iran (41).

The magnitude of MRSA detected in the current study (16.3%) is consistent with reports from various parts of the world (11.2%-17.3%) (12, 15, 36, 43). However, it is lower than the MRSA rates reported in studies from Pakistan (76%) (44), Iran (74.2%) (45), Yemen (65%) (26), different regions of Ethiopia (30%-45%) (16, 17, 35, 46), Sudan (41%) (47), and Nepal (31.2%) (48).

Interestingly, male participants were about 84% less likely to be colonized with MRSA, which aligns with findings from other parts of Ethiopia (17, 39). Similar to the patterns observed for *S. aureus* and VRSA colonization, participants with a history of ICU hospitalization were about 8 times more likely to be colonized with MRSA compared to those without such a history.

## Limitation of the study

The present study did not employ the broth dilution method for the determination of VRSA. As a result, this work should be interpreted while considering the inherent limitations associated with the use of the E-test technique for VRSA detection.

## Conclusions

The study found that the proportion of vancomycin-resistant and vancomycin-intermediate Staphylococcus aureus (VRSA and VISA) was relatively high in the study area. Importantly, a history of hospitalization in the Intensive Care Unit (ICU) and having domestic animals at home were significantly associated with VRSA colonization. Further analysis revealed that most of the VRSA isolates were resistant to aztreonam, azithromycin, clindamycin, and gentamicin. In contrast, more than 80% of the VRSA isolates were susceptible to tigecycline and doxycycline.

## Abbreviations

VRSA: Vancomycin-resistant *S. aureus*
VISA: Vancomycin intermediate *S. aureus*
HUCSH: Hawassa University Comprehensive Specialized Hospital, E-test: Epsilometer test
MRSA: Methicillin resistant *S. aureus*
MHA: Muller Hinton Agar
CLSI: Clinical Laboratory Standard Institute, ICU: Intensive care unit

## Supporting information

Not applicable

## Data Availability

All data produced in the present study are available upon reasonable request to the authors

## Acknowledgments

We would like to acknowledge nurses and laboratory scientist working at selected wards of HUCSH and teaching Microbiology laboratory respectively. We also acknowledge study participants for their willingness to take part in the study.

## Declaration

### Ethical approval and consent to participate

Ethical approval was obtained from the institutional review board (IRB) of Hawassa University College of Medicine and Health Sciences with Ref No of IRB/171/15. Written informed consent was obtained from adults participants. For minors, both consent and assent was obtained. All methods were performed in accordance with the relevant guidelines and regulations.

## Availability of data and materials

The data set used can be obtained from corresponding author.

## Authors’ contributions

**BKM:** Proposal development, laboratory work, data analysis, and drafting the original manuscript **TA:** supervision of the study, data analysis, and review of the manuscript **THM:** supervision of the laboratory work, and review of the manuscript **MMA**: proposed the study, supervision of laboratory work, data analysis and manuscript preparation

## Consent for publication

Not applicable

