## Supplementary material for "Nasal colonizing vancomycin-resistant and intermediate *Staphylococcus aureus* among admitted patients": Not applicable

Supplement table 1. Bivariate and multivariable analysis of factors associated with overall *S. aureus* colonization rate among patients admitted at HUCSH, Hawassa, Ethiopia, April to June 2023 (N=378).

| **Variables** | | **Frequency (%)** | ***S. aureus*** | | **COR(95% CI)** | ***P*-value** | **AOR (95% CI)** | ***P*-value** |
| --- | --- | --- | --- | --- | --- | --- | --- | --- |
|  |  |  | **Positive, n(%)** | **Negative, n(%)** |  |  |  |  |
| Sex | Male | 215 (56.9) | 66 (30.7) | 149 (69.4) | 2.3(1.38-3.83) | 0.001 | 3.67(1.4-9.6)* | 0.008 |
|  | Female | 163 (43.1) | 26 (16) | 137 (84) | 1 |  | 1 |  |
| Place of residence | Rural | 178 (47.1) | 42 (23.6) | 136 (76.4) | 0.93(0.57-1.48) | 0.75 |  |  |
|  | Urban | 200 (52.9) | 50 (25) | 150 (75) | 1 |  |  |  |
| Ward | Oncology | 123 (32.5) | 22 (17.8) | 101 (82.2) | 1 |  | 1 |  |
|  | Ophthalmology | 79 (20.9) | 25(31.6) | 54 (68.4) | 2.12(1.1-4.12) | 0.025 | 1.49(0.38-5.83) | 0.57 |
|  | Surgical | 60 (15.8) | 10 (16.6) | 50 (83.3) | 0.92(0.4-2.1) | 0.84 | 1.43(0.33-6.23) | 0.63 |
|  | Medical | 40 (10.6) | 12 (30) | 28 (70) | 1.97(0.87-4.46) | 0.105 | 2.03(0.52-7.83) | 0.305 |
|  | Orthopedics | 52 (13.7) | 18 (34.6) | 34 (65.4) | 2.43(1.2-5.06) | 0.018 | 1.41(0.36-5.51) | 0.62 |
|  | Neurology | 24 (6.4) | 5 (20.8) | 19 (79.2) | 1.21(0.41-3.58) | 0.733 | 0.34(0.032-3.61) | 0.37 |
| Occupation | Self-employment | 265 (70.1) | 72 (27.2) | 193 (72.8) | 1 |  |  |  |
|  | Gov’t employment | 37 (9.8) | 9 (24.3) | 28 (75.7) | 0.86(0.4-1.9) | 0.71 |  |  |
|  | House wife | 70 (18.5) | 10 (14.3) | 60 (85.7) | 0.45(0.22-0.92) | 0.029 |  |  |
|  | Other employment | 6 (1.6) | 1 (16.7) | 5 (83.3) | 0.54(0.062-4.67) | 0.57 |  |  |
| Educational status | No formal education | 131 (34.6) | 30 (22.9) | 101 (77.1) | 1 |  |  |  |
|  | 1-8 | 168 (44.4) | 45 (26.8) | 123 (73.2) | 1.23(0.72-2.1) | 0.44 |  |  |
|  | 9-12 | 54 (14.3) | 10 (18.5) | 44 (81.5) | 0.76(0.34-1.7) | 0.51 |  |  |
|  | >12 | 25 (6.6) | 7 (28) | 18 (72) | 1.31(0.5-3.43) | 0.58 |  |  |
| Marital status | Married | 189 (50) | 49 (25.9) | 140 (74.1) | 1 |  |  |  |
|  | Single | 100 (26.4) | 22 (22) | 78 (78) | 0.81(0.45-1.43) | 0.46 |  |  |
|  | Widowed | 31 (8.2) | 22 (70.9) | 9 (9.1) | 1.17(0.5-2.71) | 0.72 |  |  |
|  | Divorced | 58 (15.3) | 12(20.7) | 46 (79.3) | 0.74(0.36-1.52) | 0.42 |  |  |
| Recurrent wound | Yes | 111(29.4) | 29 (26.2) | 82 (73.8) | 1.145(0.68-1.9) | 0.602 |  |  |
|  | No | 267(70.6) | 63 (23.6) | 204 (76.4) | 1 |  |  |  |
| Current nasal infection/itching | Yes | 132 (34.9) | 33 (25) | 99 75) | 1.06(0.65-1.73) | 0.83 |  |  |
|  | No | 246 (65.1) | 59 (24) | 187 (76) | 1 |  |  |  |
| History of Hospitalization in the last 6-month | Yes | 169 (44.7) | 41 (24.3) | 128 (75.7) | 0.99(0.62-1.59) | 0.975 |  |  |
|  | No | 209 (55.3) | 51 (24.4) | 158 (75.6) | 1 |  |  |  |
| Repeated use of Antibiotics | Yes | 126 (33.3) | 35 (27.7) | 91 (72.3) | 1.32(0.81-2.15) | 0.27 |  |  |
|  | No | 252 (66.7) | 57 (22.6) | 195 (77.4) | 1 |  |  |  |
| Use of Antibiotics with physician prescription | Yes | 65 (51.6) | 26 (40) | 39(60) | 1 |  | 1 |  |
|  | No | 61(48.4) | 9 (14.8) | 52 (85.2) | 0.26(0.11-0.62) | 0.002 | 0.28(0.11-0.73)* | 0.009 |
| History of ICU Hospitalization | Yes | 67(17.7) | 21(31.3) | 46(68.7) | 1.54(0.86-2.76) | 0.143 | 3.34(1.2-9.46)* | 0.023 |
|  | No | 311(82.3) | 71(22.8) | 240(77.2) | 1 |  | 1 |  |
| Prescience of any co-morbidity | Yes | 93(24.6) | 20(21.5) | 73(78.5) | 0.78(0.446-1.37) | 0.39 |  |  |
|  | No | 285(75.4) | 72(25.3) | 213(74.7) | 1 |  |  |  |
| IV/Catheter in the body | Yes | 229(60.6) | 57(24.9) | 172(75.1) | 1.08(0.66-1.75) | 0.76 |  |  |
|  | No | 149(39.4) | 35(23.5) | 114(76.5) | 1 |  |  |  |
| Regular hand washing | Yes | 216(57.1) | 56(25.9) | 160(74.1) | 1 |  |  |  |
|  | No | 162(42.9) | 36(22.2) | 126(77.8) | 0.82(0.5-1.32) | 0.4 |  |  |
| Repeated use of public toilet | Yes | 202(53.4) | 58(28.7) | 144(71.3) | 1.68(1.04-2.73) | 0.035 | 1.96(0.75-5.14) | 0.17 |
|  | No | 176(46.6) | 34(19.3) | 142(80.7) | 1 |  | 1 |  |
| Domestic animal in home | Yes | 214(56.6) | 49(22.9) | 165(77.1) | 0.836(0.52-1.34) | 0.456 |  |  |
|  | No | 164(43.4) | 43(26.2) | 121(73.8) | 1 |  |  |  |

CI: Confidence interval, OR: Odds ratio, AOR: Adjusted odds ratio, ICU: Intensive care unit, COR: Crude odd ratio, AOR: Adjusted odd ratio

Supplement table 2. Bivariate and multivariable logistic regression of factors associated with MRSA carriage among patients admitted at HUCSH, Hawassa, Ethiopia, April to June 2023 (N=92).

| **Variables** | | **Frequency (%)** | **MRSA** | | **COR(95% CI)** | ***P*-value** | **AOR (95% CI)** | ***P*-value** |
| --- | --- | --- | --- | --- | --- | --- | --- | --- |
|  |  |  | **Positive, n (%)** | **Negative, n (%)** |  |  |  |  |
| Sex | Male | 66 (71.7) | 8 (12.1) | 58 (87.9) | 0.37(0.12-1.17) | 0.091 | 0.16(0.035-0.73)* | 0.018 |
|  | Female | 26 (28.3) | 7 (26.9) | 19 (73.1) | 1 |  | 1 |  |
| Region | Oromiya | 35 (38) | 4 (11.4) | 31 (88.6) | 1 |  |  |  |
|  | Sidama | 39 (42.4) | 7 (18) | 32 (82) | 1.69(0.45-6.37) | 0.43 |  |  |
|  | SNNPRS | 13 (14.1) | 3 (23.1) | 10 (76.9 | 2.32(0.44-12.2) | 0.32 |  |  |
|  | Amhara | 4 (4.3) | 1 (25) | 3 (75) | 2.58(0.21-31.2) | 0.45 |  |  |
|  | Addis- Ababa | 1 (1.1) | 0 (0) | 1 (100) | 1 |  |  |  |
| Place of residence | Rural | 42 (45.6) | 5 (11.9) | 37 (98.1) | 0.54(0.17-1,73) | 0.30 |  |  |
|  | Urban | 50 (54.4) | 10 (20) | 40 (80) | 1 |  |  |  |
| Occupation | Self-employment | 72 (78.3) | 11 (15.3) | 61 (84.7) | 1 |  |  |  |
|  | Government Employment | 9 (9.8) | 2 (22.2) | 7 (77.8) | 1.58(0.29-8.65) | 0.59 |  |  |
|  | House wife | 10 (10.9) | 2 (20) | 8 (80) | 1.38(0.26-7.42) | 0.7 |  |  |
|  | Other employment | 1 (1.1) | - | 1 |  | 1 |  |  |
| Educational status | No formal education | 30 (32.6) | 4 (13.3) | 26 (86.7) | 1 |  |  |  |
|  | 1-8 | 45 (48.9) | 10 (22.2) | 35 (77.8) | 0.92(0.087-9.82) | 0.95 |  |  |
|  | 9-12 | 10 (10.8) | - | 10 (100) | 1.71(0.18-15.9) | 0.64 |  |  |
|  | >12 | 7 (7.6) | 1 (14.3) | 6 (85.7) |  | 1 |  |  |
| Marital status | Married | 49 (53.3) | 6 (12.2) | 43 (87.8) | 0.69(0.12-3.98) | 0.68 |  |  |
|  | Single | 23 (25) | 6 (26.1) | 17 (73.9) | 1.76(0.29-10.45) | 0.53 |  |  |
|  | Widowed | 8 (8.9) | 1 (12.5) | 7 (87.5) | 0.71(0.054-9.5) | 0.79 |  |  |
|  | Divorced | 12 (13) | 2 (16.7) | 10 (83.3) | 1 |  |  |  |
| Ward | Oncology | 22 (23.9) | 3 (13.6) | 19 (86.4) | 1 |  |  |  |
|  | Ophthalmology | 25 (27.2) | 2 (8) | 23 (92) | 0.55(0.083-3.64) | 0.54 |  |  |
|  | Surgical | 10 (10.9) | 4 (40) | 6 (60) | 4.22(0.73-24.4) | 0.11 |  |  |
|  | Orthopedics | 18 (19.6) | 2 (11.1) | 16 (88.9) | 0.79(0.12-5.34) | 0.81 |  |  |
|  | Medical | 12 (13) | 3 (25) | 9 (75) | 2.11(0.35-12.6) | 0.41 |  |  |
|  | Neurology | 5 (5.4) | 1 (20) | 4 (80) | 1.58(0.13-19.42) | 0.72 |  |  |
| Recurrent wound | Yes | 29 (31.5) | 6 (20.7) | 23 (79.3) | 1.56(0.49-4.9) | 0.44 |  |  |
|  | No | 63 (68.5) | 9 (14.3) | 54 (85.7) | 1 |  |  |  |
| Current nasal infection/itching | Yes | 33 (35.9) | 8 (24.2) | 25 (75.8) | 2.37(0.77-7.3) | 0.13 | 1.05(0.25-4.45) | 0.95 |
|  | No | 59 (64.1) | 7 (11.9) | 52 (88.1) | 1 |  | 1 |  |
| History of Hospitalization in the last 6-month | Yes | 41 (44.6) | 7 (17.1) | 34 (82.9) | 1.11(0.36-3.36) | 0.86 |  |  |
|  | No | 51 (55.4) | 8 (15.7) | 43 (84.3) | 1 |  |  |  |
| Repeated use of Antibiotics | Yes | 35 (38) | 8 (22.8) | 27 (77.2) | 2.12(0.69-6.470 | 0.18 | 1.57(0.34-7.27) | 0.56 |
|  | No | 57 (61.9) | 7 (12.3) | 50 (87.7) | 1 |  | 1 |  |
| Use of antibiotics with physician prescription | Yes | 9 (9.8) | 3 (33.3) | 6 (66.7) | 1 |  |  |  |
|  | No | 26 (28.2) | 5 (19.2) | 21 (80.8) | 2.1(0.38-11.4) | 0.39 |  |  |
| History of ICU Hospitalization | Yes | 21 (22.8) | 8 (38.1) | 13 (61.9) | 5.63(1.73-18.2) | 0.004 | 8.06(1.72-37.6)* | 0.008 |
|  | No | 71 (77.2) | 7 (9.8) | 64 (90.1) | 1 |  | 1 |  |
| Prescience of any co-morbidity | Yes | 20 (21.7) | 3 (15) | 17 (85) | 0.88(0.22-3.49) | 0.86 |  |  |
|  | No | 72 (78.3) | 12 (16.7) | 60 (83.3) | 1 |  |  |  |
| IV/Catheter in the body | Yes | 57 (61.9) | 11 (19.3) | 46 (80.7) | 1.85(0.54-6.35) | 0.33 |  |  |
|  | No | 35 (38.1) | 4 (11.5) | 31 (88.5) | 1 |  |  |  |
| Regular hand washing | Yes | 56 (60.9) | 7 (12.5) | 49 (87.5) | 0.5(0.16-1.53) | 0.22 | 0.38(0.098-1.53) | 0.17 |
|  | No | 36 (39.1) | 8 (22.2) | 28 (77.8) | 1 |  | 1 |  |
| Repeated use of public toilet | Yes | 58 (63.1) | 10 (17.2) | 48 (82.8) | 1.21(0.37-3.88) | 0.75 |  |  |
|  | No | 34 (36.9) | 5 (14.7) | 29 (85.3) | 1 |  |  |  |
| Domestic animal in home | Yes | 49 (53.3) | 8 (16.3) | 41 (83.7) | 1.003(0.33-3.04) | 0.99 |  |  |
|  | No | 43(46.7) | 7(16.3) | 36 (83.7) | 1 |  |  |  |

MRSA: methicilin resistant *S. aureus*, CI: Confidence interval, OR: Odds ratio, AOR: Adjusted odds ratio, ICU: Intensive care unit, AOR: Adjusted odds ratio, ICU: Intensive care unit, COR: Crude odd ratio, AOR: Adjusted odd ratio
